# Strong neutralizing antibody responses to SARS-CoV-2 variants following a single vaccine dose in subjects with previous SARS-CoV-2 infection

**DOI:** 10.1101/2022.07.04.22277223

**Authors:** Nina Ekström, Anu Haveri, Anna Solastie, Camilla Virta, Pamela Österlund, Hanna Nohynek, Tuomo Nieminen, Lauri Ivaska, Paula A. Tähtinen, Johanna Lempainen, Pinja Jalkanen, Ilkka Julkunen, Arto A. Palmu, Merit Melin

**Author notes:** **Corresponding author** Nina Ekström, Finnish Institute for Health and Welfare, POB 30, 00271 Helsinki, Finland. **Funding** This study was funded by the Finnish Institute for Health and Welfare and the Academy of Finland [grant numbers 336410, 336431] and Jane and Aatos Erkko Foundation [grant number 5360-cc2fc].

## Abstract

**Background:** Previous SARS-CoV-2 infection primes the immune system and thus individuals who recovered from infection have enhanced immune responses to subsequent vaccination (hybrid immunity). However, it remains unclear how well hybrid immunity induced by severe or mild infection can cross-neutralize emerging variants. We aimed to compare the strength and breadth of antibody responses in vaccinated recovered and uninfected subjects.

**Methods:** We measured spike-specific IgG and neutralizing antibodies (NAbs) from vaccinated subjects including 320 with hybrid immunity and 20 without previous infection. From 29 subjects with a previous severe or mild infection, we also measured NAb responses against Alpha (B.1.1.7), Beta (B.1.351), Delta (B.1.617.2) and Omicron (B.1.1.529/BA.1) variants following vaccination.

**Results:** A single vaccine dose induced 2-fold higher anti-spike IgG concentrations and 3-fold higher neutralizing potency of antibodies in previously infected compared to uninfected fully vaccinated subjects. We found similar IgG concentrations in previously infected subjects after one or two vaccine doses. NAb titers were higher in subjects with severe compared to those with mild infection. This difference remained after vaccination with sequentially decreasing titers against Alpha, Beta, Delta, and Omicron variants.

**Conclusions:** Hybrid immunity induced strong IgG responses, particularly after severe infection. However, the NAb titers were low against heterologous variants, especially against Omicron.

## Background

Infection with severe acute respiratory syndrome coronavirus 2 (SARS-CoV-2) induces antibodies to the viral spike glycoprotein (S), which is also the target of coronavirus disease 2019 (COVID-19) vaccines. The generation of neutralizing antibodies (NAbs) that specifically target the receptor-binding domain (RBD) of the S protein is considered essential in controlling SARS-CoV-2 infection. We and others have previously shown that circulating antibodies gradually decrease following wild-type (WT) infection but that NAbs are sustained at a detectable level for at least up to 15 months [1, 2]. However, antibody-mediated immunity induced by infection with the ancestral virus is reduced against variants with immune escape mutations, as only part of the NAbs can bind to the RBD of these variants [3, 4]. The Omicron variant carries several mutations in the RBD [5, 6] and has rapidly given rise to major epidemic waves worldwide since late 2021, also causing breakthrough infections in vaccinated individuals.

COVID-19 vaccination after recovery from SARS-CoV-2 infection (hybrid immunity) has been reported to induce comparable or higher S-specific antibody levels and NAb titers than in twice vaccinated SARS-CoV-2 naïve individuals [7-12]. In addition, vaccination has been shown to elicit immunity with broader specificity and increase the neutralization potency against SARS-CoV-2 variants in previously infected individuals [13, 14]. Previous studies have shown no increase in circulating antibodies, neutralizing titers or antigen-specific memory B cells after more than one dose of vaccine in those with previous infection [8, 15, 16]. Hybrid immunity-induced antibody concentrations and NAbs have been shown to decline with time but remain at a higher level than in uninfected vaccinated individuals for at least three months [8, 11, 17]. Furthermore, hybrid immunity has been associated with a somewhat lower risk of reinfection and hospitalization compared to immunity induced solely by previous infection [18-20]. To better understand the level of protection hybrid immunity provides against different SARS-CoV-2 variants, including Omicron (B.1.1.529), we compared the strength and breadth of IgG and NAb responses induced by hybrid immunity to vaccination or infection alone and assessed hybrid immunity induced by severe and mild SARS-CoV-2 infection.

## Methods

### Study design

This was an observational study assessing immune responses induced by SARS-CoV-2 infection and vaccination. Vaccinations were administered according to the Finnish national COVID-19 vaccination programme starting in December 2020. We collected blood samples after infection and vaccination (Figure 1), separated the specimens by centrifugation and stored aliquoted serum at - 20°C or -70°C. We defined the COVID-19 severity as severe or mild. Severe infection was defined as a laboratory-confirmed COVID-19 requiring hospital treatment based on data collected from the hospital discharge register (Care Register for Health Care), and mild infection as laboratory-confirmed COVID-19 without documentation of hospital treatment. We collected demographics, clinical characteristics, and COVID-19 vaccination history of the participants from the National Infectious Disease Register and the National Vaccination Register (Table 1). The study protocol of the serological population study of the coronavirus epidemic was approved by the ethical committee of the Hospital District of Helsinki and Uusimaa [HUS/1137/2020]. The study protocol of the study COVID-19 infections in hospital personnel [21] was approved by the ethical committee of the Southwest Finland health district [ETMK 19/1801/2020] and by the Finnish Medicines Agency Fimea as the regulatory authority [European Union drug regulation authority’s clinical trials database code 2021-004419-14]. Written informed consent was obtained from all participants before sample collection.

**Figure 1.**
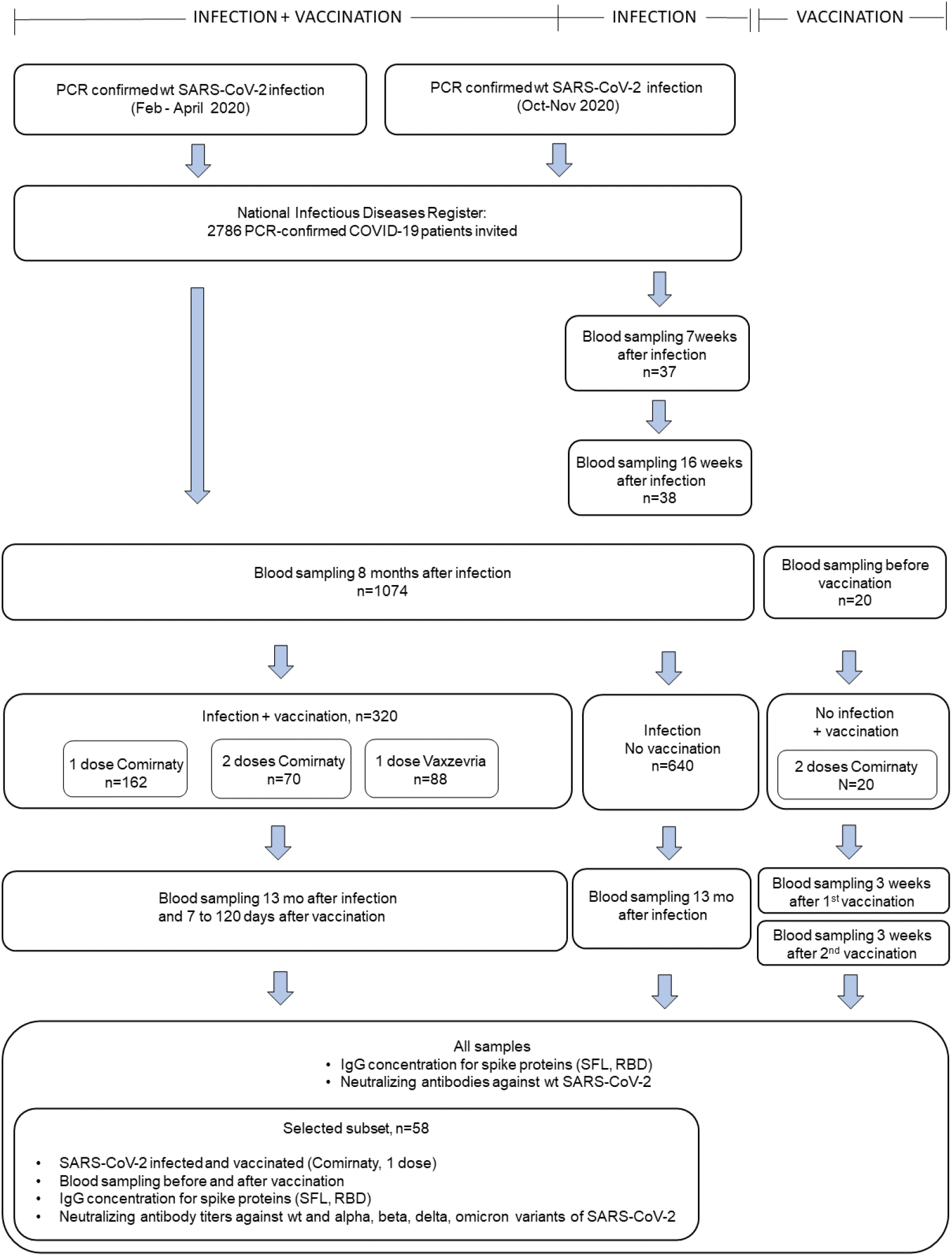
Selection of study subjects with or without a previous SARS-CoV-2 infection and with or without a subsequent SARS-CoV-2 vaccination, and selection of serum samples for the determination of spike protein specific IgG antibody concentration and neutralizing antibodies.

**Table 1.** Characteristics of the participants in the study subgroups and timing of vaccination and serum sampling

| Infection and vaccination status,<br>days from vaccination or infection | n | Days from<br>infection to<br>vaccination,<br>median (range) |  | Age,<br>median (range) |  | Severe disease<br>n (%) |  | Female<br>(%) |
| --- | --- | --- | --- | --- | --- | --- | --- | --- |
| **Infected, Comirnaty, one dose** |  |  |  |  |  |  |  |  |
| 7-30 | 102 | 369 | (342-424) | 62 | (26-79) | 23 | (23) | 58 |
| 31-60 | 50 | 353 | (312-419) | 73 | (30-96) | 11 | (22) | 64 |
| 61-90 | 10 | 310 | (299-334) | 77 | (29-90) | 6 | (60) | 60 |
| **Subset selected for NAb titration** |  |  |  |  |  |  |  |  |
| 22-85 (severe infection) | 15 | 338 | (307-377) | 77 | (69-97) | 15 | (100) | 33 |
| 22-83 (mild infection) | 14 | 355 | (299-415) | 52 | (28-76) | 0 | (0) | 64 |
| **Infected, Vaxzevria, one dose** |  |  |  |  |  |  |  |  |
| 7-30 | 29 | 372 | (349-399) | 67 | (62-73) | 13 | (45) | 38 |
| 31-60 | 37 | 340 | (313-360) | 57 | (28-67) | 13 | (35) | 62 |
| 61-90 | 22 | 322 | (290-351) | 56 | (38-67) | 6 | (27) | 73 |
| **Infected, Comirnaty, two doses** |  |  |  |  |  |  |  |  |
| 7-30<sup>#</sup> | 12 | 292 | (267-313) | 52 | (27-66) | 1 | (8) | 92 |
| 61-90<sup>*</sup> | 29 | 280 | (261-293) | 52 | (26-62) | 2 | (7) | 83 |
| 91-120<sup>*</sup> | 30 | 270 | (250-314) | 56 | (25-91) | 4 | (13) | 90 |
| **Infected, not vaccinated** |  |  |  |  |  |  |  |  |
| 14-60 | 37 | - | - | 47 | (23-80) | 3 | (8) | 59 |
| 90-148 | 38 | - | - | 47 | (23-80) | 3 | (8) | 63 |
| 362-448 | 640 | - | - | 48 | (18-81) | 78 | (12) | 59 |
| **Uninfected, Comirnaty** |  |  |  |  |  |  |  |  |
| 14-30 (after one dose) | 20 | - | - | 42 | (25-61) | - | - | 90 |
| 14-30 (after two doses) | 20 | - | - | 42 | (25-61) | - | - | 90 |

<sup>#</sup>1<sup>st</sup> and 2<sup>nd</sup> vaccine at a median of 84 days apart (range 49-88)<sup>\*</sup>1<sup>st</sup> and 2<sup>nd</sup> vaccine at a median of 21 days apart (range 19-28)

#### Samples from infected, vaccinated subjects

We identified 2586 subjects ≥18 years of age with PCR-confirmed SARS-CoV-2 infection between February and April 2020 in the National Infectious Disease Register and invited them to participate [1]. The participants were infected when the B, B.1 and B.1.1 lineage viruses (hereafter referred as wildtype, WT) were introduced into Finland [22]. We collected a blood sample for determination of SARS-CoV-2 specific serum antibodies from 1074 subjects at 8 months (median 7.7, range 6.1 to 11.4) and 13 months (median 12.9, range 11.8 to 17.3) after SARS-CoV-2 infection. In this study, we included participants who had received one dose of COVID-19 vaccine (BNT162b2, Comirnaty, Pfizer-BioNTech, n=162 or ChAdOx1, Vaxzevria, AstraZeneca, n=88) 7 to 90 days or two doses of Comirnaty (n=70) 7 to 120 days before the 13-month sampling (Figure 1, Table1).

We selected paired serum samples from 30/55 of the study participants with a previous SARS-CoV-2 infection followed by one dose of Comirnaty 22 to 90 days before sampling for the determination of NAb titers against WT virus, and four variants; Alpha (B.1.1.7), Beta (B.1.351), Delta (B.1.617.2) and Omicron (B.1.1.529/BA.1)(Figure 1). We first selected 15/15 subjects with a sample taken 59 to 90 days after vaccination; 8/15 of these subjects had previous severe disease. We then selected 15/40 subjects with sample taken 21 to 30 days after vaccination by matching with disease severity, age and gender to the first group. One subject with mild disease was afterwards excluded due to administration of two doses of Comirnaty before sampling.

#### Samples from infected, not vaccinated subjects

We collected serum samples from 640 subjects ≥18 years of age 13 months after PCR-confirmed WT SARS-CoV-2 infection between February and April 2020 with no documentation of COVID-19 vaccination before sampling [1] (Figure 1, Table 1). We excluded subjects with >30% increase in IgG antibodies to nucleoprotein or spike protein between samples taken at 8 and 13 months after infection. We additionally collected serum samples from 38 subjects with no COVID-19 vaccination history and with a previous WT SARS-CoV-2 infection (between October and November 2020) at a median of 51 days (range 19 to 59) and 118 days (range 90 to 148) following infection (Figure 1, Table1).

#### Samples from vaccinated, uninfected subjects

Serum samples from subjects with no history of previous SARS-CoV-2 infection and who had received two doses of COVID-19 vaccine (Comirnaty) were obtained from Turku University Hospital (TUH, Turku, Finland). Blood samples from 20 subjects were collected at a median of 21 days (range 18-21) after the first vaccine dose and 27 (range 18-29) days after the second vaccine dose (at 39-50 days after the first vaccine dose).

### Laboratory methods

#### SARS-CoV-2 fluorescent multiplex immunoassay

We measured the concentration of serum IgG antibodies to WT spike glycoprotein of SARS-CoV-2 (S-IgG; full-length spike protein, SFL-IgG, and receptor binding domain of spike protein, RBD-IgG) with an in-house fluorescent multiplex immunoassay (FMIA) as previously described [23]. Antibody concentrations are given as WHO international binding antibody units (BAU)/ml [24]. IgG SARS-CoV2 FMIA is an accredited assay at the Expert Microbiology Unit of the Finnish Institute of Health and Welfare which is a testing laboratory T077 accredited by FINAS Finnish Accreditation Service, accreditation requirement SFS-EN ISO/IEC 17025.

#### SARS-CoV-2 microneutralization test (MNT)

We performed a cytopathic effect-based, live-virus microneutralization test (MNT) as previously described [1, 3, 25]. All samples were screened with WT virus (B lineage, Supplementary Table 1) for NAb positivity. NAb titers for a selected subset of samples were analyzed with four SARS-CoV-2 variants isolated in Finland during 2021 (Supplementary Table 1: Alpha (B.1.1.7), Beta (B.1.351), Delta (B.1.617.2) and Omicron (B.1.1.529/BA.1)). WT virus isolation and propagation were performed in African green monkey kidney epithelial (VeroE6) cells [25]. All variant viruses were isolated and propagated in VeroE6-TMPRSS2-H10 cells [26] and further propagated in VeroE6 cells for MNT. A tissue culture infectious dose 50% (TCID_50_) /ml titer was determined with an end-point dilution assay with the same incubation times as MNT for all viruses to achieve a comparable virus concentration (100 TCID_50_ per well) among the different virus strains.

Results were expressed as MNT titers corresponding to the reciprocal of the serum dilution that inhibited 50% of SARS-CoV-2 infection. A MNT titer ≥6 was considered positive, 4 borderline, and <4 negative. Borderline values were confirmed with biological repeats. For comparison, a titer of 192, 96, 8, 32 and <4 was measured for the WHO International Standard (NIBSC 20/136) [24] for the WT virus, Alpha, Beta, Delta and Omicron BA.1 variant, respectively.

The neutralizing potency (neutralizing activity adjusted for IgG antibody concentration) of serum was calculated as the ratio of neutralizing activity (MNT titer) to anti-spike IgG antibody concentration (BAU/ml) and was determined for samples taken following hybrid immunity (selected subgroup, n=29) and following two doses of vaccine from uninfected subjects (n=20).

### Statistical methods

Our aim was to assess antibody levels induced in previously infected subjects following vaccination, compared to previously uninfected fully vaccinated and previously infected unvaccinated subjects. We calculated the geometric mean concentrations (GMCs) and titers (GMTs) with 95% confidence intervals (CI) for IgG and NAbs, respectively. For GMT calculation, MNT titers <4 were assigned a titer value of 2. We assessed the statistical differences in antibody levels and neutralizing potencies between the groups using the Mann Whitney test and differences in neutralizing antibody titers between viruses with Wilcoxon rank sum test. We set the statistical significance level of difference to *P*<0.05. We performed the statistical analyses using GraphPad (v9) and R (v4.0.4) with Rstudio (v1.4.1106).

## Results

### Spike-specific and neutralizing antibodies following hybrid immunity and vaccination alone

We measured spike-specific antibodies in previously SARS-CoV-2 infected subjects following one dose of COVID-19 vaccine. We found that before vaccination at eight months after infection 97% of the subjects were positive for S-IgG and 89% had NAbs against WT SARS-CoV-2. A single vaccine dose at a median of 351 days (range 250 to 424) after infection induced >20-fold higher IgG concentrations in previously infected compared to uninfected subjects (Table 2, Figure 2). We additionally found that following hybrid immunity induced by mild infection the mean S-IgG concentrations were notably higher (>15-fold) compared to S-IgG concentrations after mild infection alone (Figure 3).

**Table 2.** Geometric mean anti-SARS-CoV-2 spike protein IgG concentrations GMC (95% CI) expressed as binding antibody units/ml for wild-type (WT) spike proteins receptor binding domain of spike (RBD) and full-length spike protein (SFL), percentage of positive samples for spike protein IgG (S-IgG) and neutralizing antibodies (NAb) in vaccinated and unvaccinated subjects with a previous SARS-CoV-2 infection and in uninfected vaccinated subjects measured from samples taken after SARS-CoV-2 infection, stratified by days of serum sample collection since last vaccination. NAbs were measured against WT SARS-CoV-2 virus.

| Infection and vaccination status,<br>Days from vaccination or<br>infection | GMC (95% CI) |  |  |  |  | % positive |  |
| --- | --- | --- | --- | --- | --- | --- | --- |
|  | n | RDB-IgG |  | SFL-IgG |  | S-IgG | NAb |
| **Infected, Comirnaty one dose** |  |  |  |  |  |  |  |
| 7-30 | 102 | 120 | (94-154) | 238 | (189-300) | 100 | 98 |
| 31-60 | 50 | 101 | (75-137) | 187 | (137-256) | 100 | 98 |
| 61-90 | 10 | 61 | (20-186) | 155 | (71-336) | 100 | 100 |
| **Infected, Vaxzevria one dose** |  |  |  |  |  |  |  |
| 7-30 | 29 | 78 | (53-115) | 138 | (95-200) | 100 | 100 |
| 31-60 | 37 | 44 | (27-74) | 68 | (40-117) | 100 | 97 |
| 61-90 | 22 | 40 | (24-65) | 51 | (30-89) | 100 | 95 |
| **Infected, Comirnaty, two doses** |  |  |  |  |  |  |  |
| 7-30<sup>#</sup> | 12 | 146 | (93-229) | 253 | (168-382) | 100 | 100 |
| 61-90<sup>§</sup> | 29 | 82 | (59-115) | 137 | (100-187) | 100 | 100 |
| 91-120<sup>§</sup> | 30 | 76 | (60-96) | 112 | (87-146) | 100 | 100 |
| **Infected, not vaccinated** |  |  |  |  |  |  |  |
| 14-60 | 37 | 2.5 | (1.0-5.9) | 2.8 | (1.3-6.0) | 89 | 92 |
| 90-148 | 38 | 1.7 | (0.79-3.5) | 2.1 | (1.1-4.2) | 89 | 89 |
| 362-448 | 640 | 1.7 | (1.6-1.9) | 2.3 | (2.1-2.5) | 97 | 90 |
| **Not infected, Comirnaty** |  |  |  |  |  |  |  |
| 14-30 (after one dose) | 20 | 3.8 | (2.1-7.1) | 9.2 | (5.7-15) | 100 | 100 |
| 14-30 (after two doses) | 20 | 63 | (42-96) | 95 | (64-142) | 100 | 100 |

<sup>#</sup>1<sup>st</sup> and 2<sup>nd</sup> vaccine at a median of 84 days apart (range 49-88)
<sup>§</sup>1<sup>st</sup> and 2<sup>nd</sup> vaccine at a median of 21 days apart (range 19-28)

**Figure 2.**
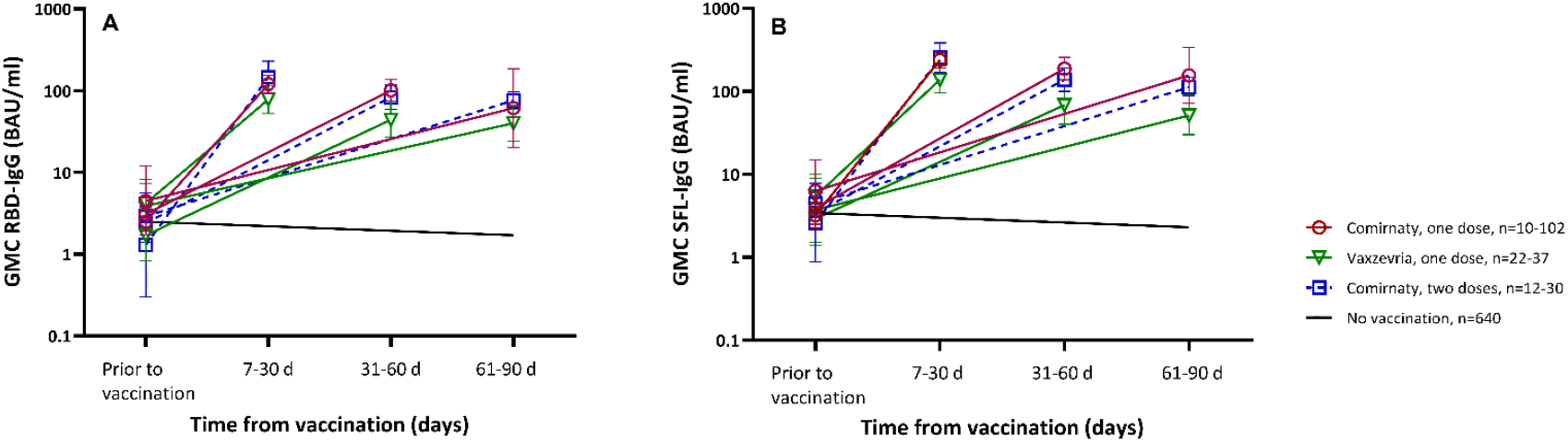
SARS-CoV-2 spike specific IgG antibody concentrations for the receptor binding domain, RBD (A) and full-length spike protein, SFL (B) expressed as binding antibody units BAU/ml (GMC, 95% CI) for samples taken prior to the first vaccine dose and 7 to 30, 31 to 60 and 61 to 90 days after the last vaccine dose (Comirnaty or Vaxzevria). Samples collected at different time points were not collected from the same subjects.

**Figure 3.**
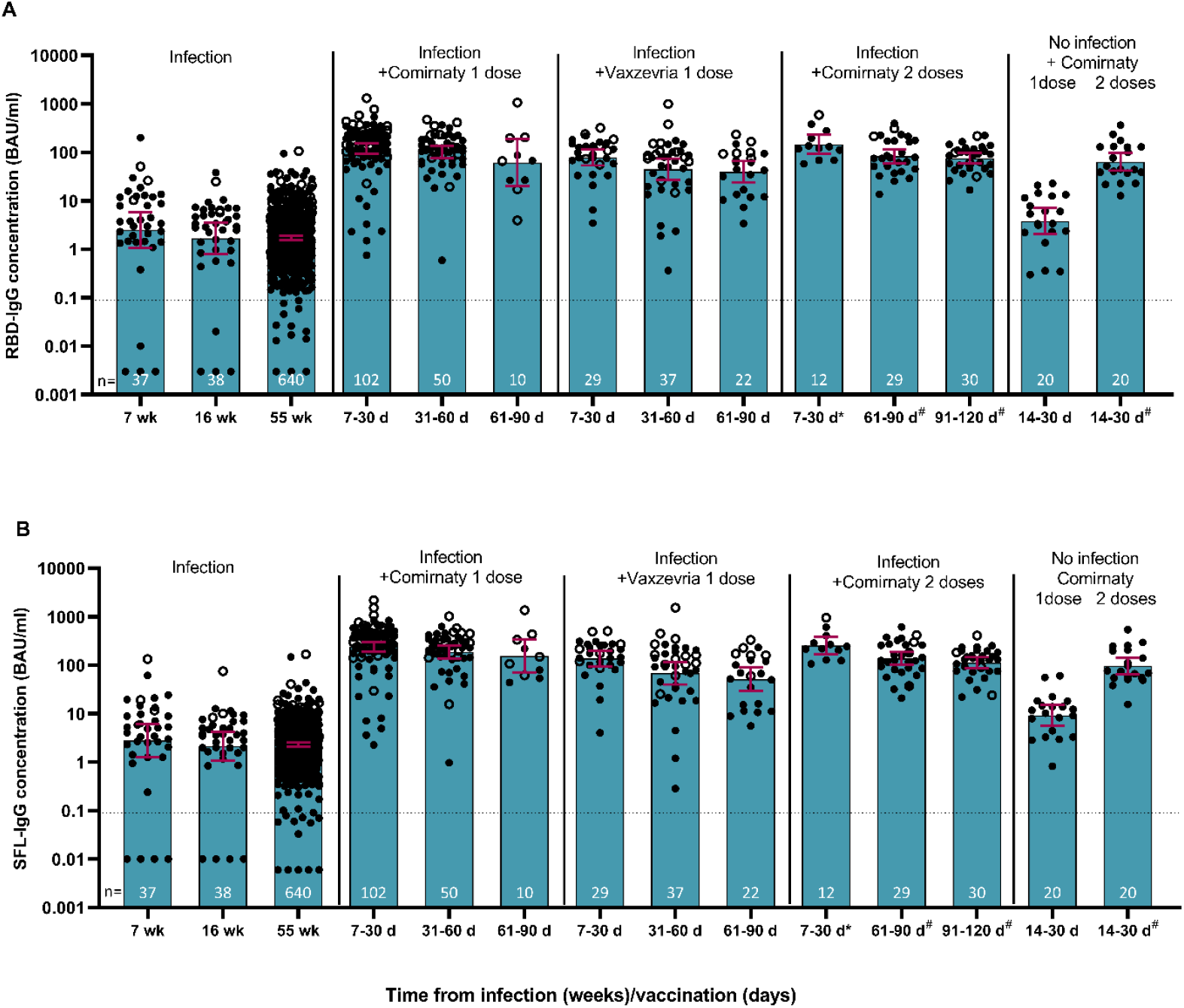
SARS-CoV-2 spike specific IgG antibody concentrations for the receptor binding domain, RBD (A) and SFL-full-length spike protein, SFL (B) expressed as binding antibody units BAU/ml (GMC, 95% CI) in subjects with or without a previous SARS-CoV-2 infection and after one or two doses of COVID-19 vaccine (Comirnaty or Vaxzevria). Samples from subjects with previous severe infection are shown by open labels. Two vaccine doses were administered either with a short dosing interval^#^ (median of 21 days apart (range 19-28) or a long dosing interval* (median of 84 days apart, range 49-88 days).

We compared the antibody concentrations in previously infected subjects after one vaccine dose (Comirnaty or Vaxzevria) to uninfected subjects after two doses of Comirnaty. We found 2-fold higher S-IgG concentrations after infection followed by one dose of Comirnaty compared to two doses of Comirnaty alone (*P*<0.001, Table 2, Figure 3). In subjects receiving Vaxzevria following infection we found similar or slightly higher S-IgG concentrations compared to two doses of vaccine (Comirnaty) alone (P= 0.237 and 0.04, for RBD- and SFL-IgG, respectively) (Table 2, Figure 3). A high percentage of subjects with hybrid immunity (98%) and twice vaccinated subjects (100%) had NAbs against WT SARS-CoV-2 (Table 2). When we compared the neutralizing potency of the antibodies targeting the spike protein we found, however, that nearly 3-fold higher neutralizing potency of antibodies was reached following hybrid immunity compared to two doses of vaccine alone (Supplementary Figure 1). At 90 days following hybrid immunity the mean antibody levels had decreased, but 97% of the subjects still had NAbs (Table 2, Figure 3).

We assessed the effect of a second vaccine dose on hybrid immunity by measuring IgG concentrations and NAbs after two doses of Comirnaty. The second dose was administered either with a short (median 21 days, range 19-28, n=59) or a long (median 84 days, range 49-88, n=12) dosing interval at a median of 277 days after infection (Table 1). We found comparable S-IgG concentrations after the first and second dose and with either a short or long dosing interval, suggesting that in hybrid immunity the second vaccine dose did not further enhance S-IgG levels (P>0.05, Table 2, Figure 2). To take into account the larger proportion of subjects with a severe disease in the one compared to two dose group, we included only subjects with a mild disease in the comparison. In line with high antibody levels induced by hybrid immunity, the proportion of subjects with NAbs was high after one (98%) and two doses of the vaccine (100%) (Table 2).

### Neutralization of SARS-CoV-2 variants after hybrid immunity

A subset of participants with a previous severe (n=15) or mild (n=14) WT SARS-CoV-2 infection were selected for NAb titration (Figure 1, Table 1). Before vaccination and at eight months after WT infection, we measured higher mean IgG concentrations and NAb titers in subjects with a severe infection compared to those with a mild infection (Table 3, Figure 4). All subjects with previous severe infection had NAbs for WT virus, Alpha and Delta variants, but the proportion of subjects with NAbs were lower against Beta (80%) and Omicron BA.1 (33%) variants. The proportions of subjects with NAbs in the mild infection group were overall lower, reaching 79%, 50%, 21% and 43% for WT virus, Alpha, Beta and Delta variants, respectively. None of the subjects with a mild infection had NAbs against the Omicron BA.1 variant eight months after infection (Table 3, Figure 4).

**Table 3.** Geometric mean titers, GMT (95% CI) of neutralizing antibody (NAb) titers to wild-type (WT) virus, Alpha (B.1.1.7), Beta (B.1.351), Delta (B.1.617.2) and Omicron (B.1.1.529/BA.1) variants, and IgG geometric mean concentrations, GMC (95% CI) expressed as binding antibody units/ml for WT SARS-CoV-2 spike proteins (RBD and SFL) eight months after severe (n=15) or mild (n=14) infection and 1 to 3 months after one dose of COVID-19 vaccine (Comirnaty) administered one year after the infection. Geometric mean fold changes (GMFC) are additionally presented for both NAb and IgG data.

|  | Severe disease n=15 |  |  | Mild disease n=14 |  |  |
| --- | --- | --- | --- | --- | --- | --- |
|  | 8 months after infection<br>GMT (95% CI) | 1 to 3 months after vaccination<br>GMT (95% CI) | GMFC | 8 months after infection<br>GMT (95% CI) | 1 to 3 months after vaccination<br>GMT (95% CI) | GMFC |
| **WT** | 44 (24-78) | 1500 (660-3400) | 34 | 6.9 (3.8-12) | 180 (57-600) | 27 |
| **Alpha** | 23 (15-37) | 1100 (460-2400) | 46 | 4.4 (2.6-7.4) | 100 (29-360) | 23 |
| **Beta** | 11 (5.7-21) | 360 (150-870) | 33 | 2.6 (1.8-3.8) | 53 (16-180) | 20 |
| **Delta** | 21 (13-35) | 630 (280-1400) | 30 | 3.7 (2.3-5.8) | 75 (22-260) | 21 |
| **Omicron** | 2.7 (2.1-3.5) | 110 (46-240) | 39 | 2.0 (2.0-2.0) | 17 (7.4-37) | 8.3 |
|  | GMC (95% CI) | GMC (95% CI) | GMFC | GMC (95% CI) | GMC (95% CI) | GMFC |
| **WT RBD-IgG** | 15 (6.8-34) | 154 (68-350) | 10 | 0.51 (0.10-2.6) | 41 (19-90) | 83 |
| **WT SFL-IgG** | 13 (8.3-21) | 340 (200-570) | 26 | 0.90 (0.21-3.8) | 84 (40-170) | 94 |

**Figure 4.**
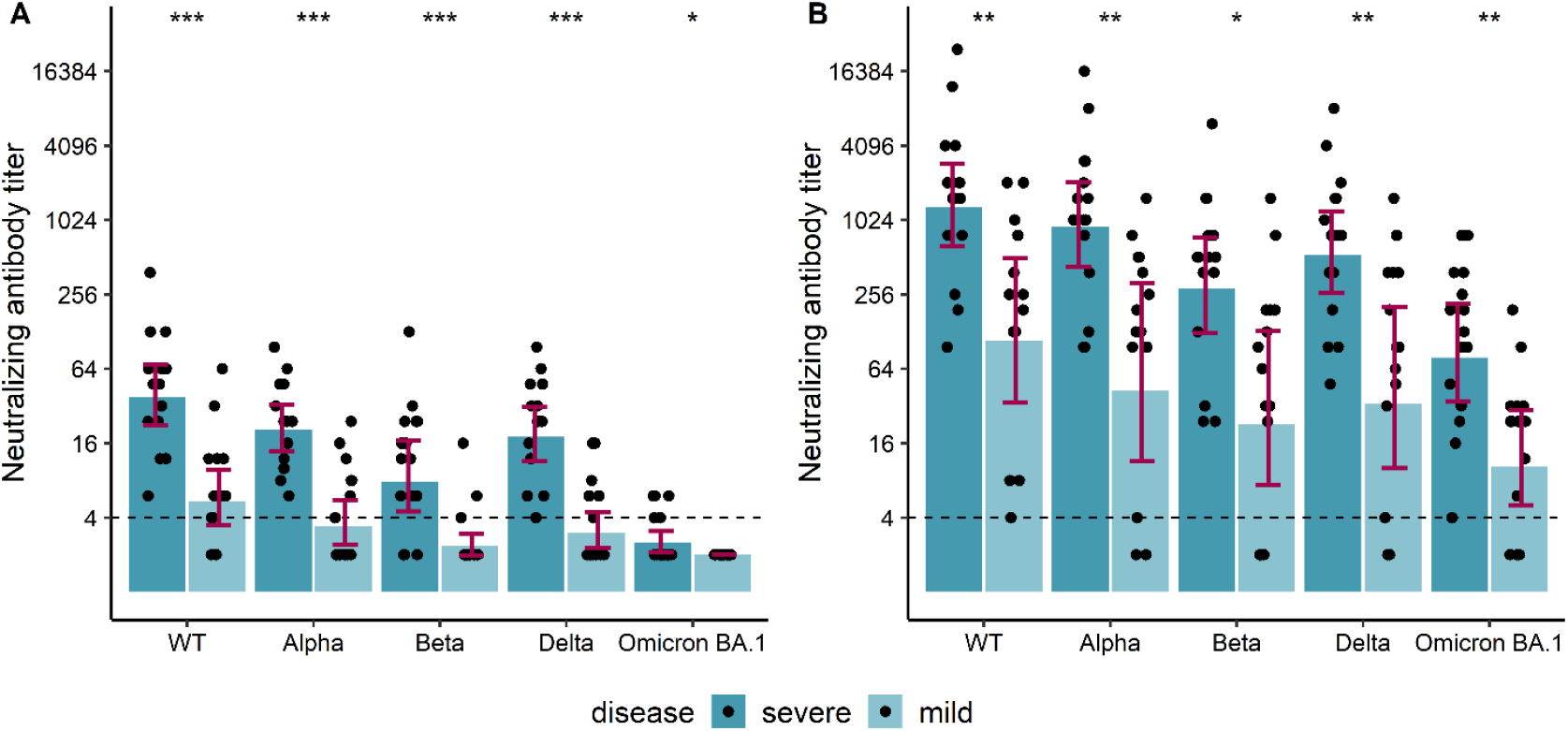
Neutralizing antibody titers to wild-type (WT) virus, Alpha (B.1.1.7), Beta (B.1.351), Delta (B.1.617.2) and Omicron (B.1.1.529/BA.1) variants in severe (n=15) or mild (n=14) infection groups at eight months after infection and before vaccination (A) and 1 to 3 months after one dose of COVID-19 vaccine (Comirnaty) administered one year after infection (B). The height of the bars represents medians and whiskers the interquartile range within infection group. Dashed line marks threshold for positive result (titer 4). Statistically significant differences between groups of mild and severe infection are indicated with asterisks (Wilcoxon rank-sum test) * = *P*<.05, ** = *P*<.01, *** = *P*<.001.

One dose of Comirnaty among the previously infected subjects induced very high mean IgG concentrations and NAb titers compared to the levels measured before vaccination (Table 3, Figure 4). We observed the highest mean NAb titers to the WT virus, and reduced titers sequentially against Alpha, Delta, Beta and Omicron BA.1 variants. Mean NAb titer fold changes ranged between 30-46 and 8-27, depending on the virus strain, for the severe and mild infection groups, respectively, suggesting that hybrid immunity is more enhanced after a severe compared to a mild infection. Subjects with a previous severe infection had NAbs to all virus strains analyzed (Figure 4). In the mild infection group, one dose of Comirnaty induced NAbs against WT virus in 100% of the subjects, and positive or borderline positive titers were measured in a total of 86%, 79%, 86% and 79% of the subjects for Alpha, Beta, Delta and Omicron BA.1 variants, respectively. The higher level of IgG concentrations and NAb titers seen in subjects with a previous severe compared to mild infection remained also after hybrid immunity (Table 3, Figure 4).

The NAb titers of three subjects within the mild infection group differed notably from the rest of the group. None of them had detectable NAbs or IgG eight months after infection. After one dose of Comirnaty, the NAb titers for WT virus were detectable but markedly lower compared to the other subjects. One of the three subjects had borderline positive antibody titers against Alpha and Delta variants, otherwise the NAb titers against variants were below the detection limit (Figure 4).

## Discussion

Our study showed that a single COVID-19 vaccine dose induced strong SARS-CoV-2 spike specific IgG and NAb responses in subjects with a previous infection with 2-fold higher IgG levels compared to previously SARS-CoV-2 naïve twice vaccinated subjects. Additionally, we found that a second vaccine dose did not further enhance the IgG antibody response induced by hybrid immunity and that a longer dosing interval (49 to 88 days) did not improve antibody response after the second dose. In two age- and gender-matched subsets of subjects with a previous severe or mild infection, we found that hybrid immunity is more enhanced after severe infection, as higher fold-increases in NAb titers were seen in subjects with severe infection following vaccination. We also found that in previously infected subjects a single vaccine dose induced NAbs with higher neutralizing potency compared to vaccination only. However, even with hybrid immunity, NAb titers remained reduced against heterologous variants, especially Omicron BA.1.

In line with our findings, other studies have also reported that in previously infected individuals vaccination induced strong antibody responses [27-31], and comparable or superior responses compared to SARS-CoV-2 naïve individuals vaccinated with two doses [7, 8, 10, 16, 32]. We have previously also shown that a single vaccine dose three to six months following WT, Alpha or Beta infection induced S-IgG concentrations comparable to the levels following a third COVID-19 vaccine dose in previously uninfected individuals, whereas cross-reactivity of NAbs against different variants appeared to be even more enhanced [3]. Bates and coworkers found that even a mild breakthrough infection caused by the Delta variant substantially boosted humoral immunity induced by vaccination and improved variant cross-neutralization [33]. The same authors also found that antigen exposure from natural infection enhanced the magnitude and breadth of the antibody response similarly regardless of whether the exposure occurred before the vaccination (hybrid immunity) or after the breakthrough infection [14]. This finding was also reported by Walls and coworkers who showed that NAb responses were comparable in breakthrough cases, hybrid immunity and in uninfected individuals vaccinated with three doses [34].

Our finding that a second vaccine dose after infection did not further increase antibody concentrations is in line with previous immunogenicity studies with dosing intervals of two to three weeks [8, 15, 35]. Moreover, a previous study conducted in Israel during the surge of the Delta variant in 2021 found that there was no significant difference in vaccine effectiveness against reinfection after one or two doses of the vaccine [18]. However, the study of Muecksch and coworkers showed that despite similar antibody concentrations after two or three doses of COVID-19 vaccine the neutralizing potency and breadth of antibodies was increased after the third compared to the second vaccine dose in previously uninfected subjects [36]. Their data indicated that the third exposure to SARS-CoV-2 antigen expanded persisting clones of memory B cells expressing more potent and broader antibodies. Further, the study of Wratil and coworkers recently reported that timely spaced, second vaccination nine months after the infection further increased neutralization capacity against different variants such as Omicron, suggesting that a longer dosing interval may be needed for optimal maturation of immunity and for more efficient cross-variant neutralization [37]. Similarly, Miyamoto and coworkers found that a longer interval between the vaccination and breakthrough infection was favorable for better antibody responses [38]. These findings suggest that maturation of the B cell response takes place during the period of several months following infection.

In this study, the NAb titer pattern was similar to that seen in our previous studies [1, 3] showing the highest titers for WT virus, and sequentially decreasing titers against Alpha, Delta, Beta and Omicron BA.1 variants. NAb titers against the Omicron BA.1 variant were the lowest suggesting reduced cross-protection against this recently circulated virus variant. None of the subjects with a mild WT infection had detectable NAbs against the Omicron BA.1 variant eight months after the infection. We have also previously found that 36/37 subjects with a recent, mild non-Omicron infection lacked NAbs against the Omicron BA.1 variant [3]. Recent, not yet peer reviewed studies have reported that Omicron infection induces stronger immune responses in previously vaccinated compared to naïve subjects [39], and that Omicron infection elicits NAbs that can cross-react with other sub-lineages of Omicron and other variants in individuals with hybrid immunity [40]. These findings emphasize the benefits of hybrid immunity and the need to vaccinate previously infected subjects to generate cross-reactive NAbs against different variants.

COVID-19 vaccination has been shown to induce lower antibody responses among individuals aged 65 and older [3, 9, 41, 42]. The relationship between age and antibody responses after infection is, however, more complex due to the greater overall disease severity among the elderly and higher antibody responses observed after a severe than a mild infection. Similarly, in this study the median age of those with a severe infection was higher than of those with a mild infection.

Those with a severe infection had markedly higher NAb and IgG levels eight months after WT infection and the NAb titers against different variants remained at a significantly higher level up to 13 months following a severe compared to mild WT infection [1]. We have previously shown that individuals ≥60 years of age with a mild infection had similar S-IgG levels and an equally high proportion of them had NAbs against WT virus compared to younger individuals with a mild infection, suggesting that a more severe disease leads to stronger humoral immune response [1]. In this study we found that this difference persisted after a single dose of COVID-19 vaccination, suggesting enhanced duration of immunity and improved protection against emerging variants for those having experienced a severe infection.

Our study had some limitations. As this study was not designed to assess the kinetics and durability of humoral immune responses, we did not collect follow-up samples from the same individuals after vaccination. Also, the study design did not allow us to compare the effect of different dosing intervals on hybrid immunity. Due to resource restrictions with the laborious live-virus neutralization assay, we did not assess NAb titers against variants in previously infected subjects after two doses of vaccine and did not assess antibody neutralizing potencies induced by hybrid immunity after one compared to two doses of the vaccine.

In summary, we found that a single dose of COVID-19 vaccine given nine to twelve months after an infection strongly enhanced spike-specific IgG and NAb levels and neutralizing potency in previously WT SARS-CoV-2 infected subjects. NAb titers in previously infected subjects were higher in subjects with a past severe than a mild disease and this difference remained after vaccination. The overall lowest NAb titers against Omicron BA.1 suggest reduced cross-protection against this variant in previously WT SARS-CoV-2 infected subjects with one dose of the vaccine.

Our data supports the importance of vaccinating both uninfected and previously infected individuals to elicit cross-variant neutralizing antibodies. Further studies are needed to elucidate whether three timely spaced encounters with the SARS-CoV-2 spike antigen would be required to combat Omicron and emerging variants more efficiently.

## Data Availability

All data produced in the present study are available upon reasonable request to the authors

## Acknowledgements

We thank all the subjects who participated in the study. We also thank Elina Isosaari, Esa Ruokokoski, Juha Oksanen, Dennis Ahlfors, Timo Koskenniemi, Mikko Aura, Katja Lind, Hanna Valtonen, Heidi Hemmilä, Marja-Liisa Ollonen, Larissa Laine, Tiina Sihvonen, Raisa Hanninen, Johanna Rintamäki, Leena Saarinen, Marja Suorsa, Camilla Virta, Lotta Hagberg, Mervi Lasander, Marja Leinonen, Hanna K. Valtonen, Jenni Krogell, Arja Rytkönen, Oona Liedes, Saimi Vara, Joni Niemi, Susanna Celik, Maila Kyrölä, Päivi Siren and Timothée Dub.

We gratefully acknowledge the authors and their respective laboratories, who analyzed and submitted the sequences to GISAID’s EpiCoV™ and GenBank® Database.

