## Supplemental material for "Strong neutralizing antibody responses to SARS-CoV-2 variants following a single vaccine dose in subjects with previous SARS-CoV-2 infection"

**Supplementary material**

**Supplementary Table 1.** Details of the SARS-CoV-2 virus strains used in live-virus microneutralization test

| WHO label | Pango lineage | Isolate | GISAID accession ID | GenBank accession ID |
|---|---|---|---|---|
| **Wild-type** | B | hCoV-19/Finland/1/2020 | EPI_ISL_407079 | MZ934691 |
| **Alpha** | B.1.1.7 | hCoV-19/Finland/THL-202102301/2021 | EPI_ISL_2590786 | MZ944886 |
| **Beta** | B.1.351 | hCoV-19/Finland/THL-202101018/2021 | EPI_ISL_3471851 | MZ944846 |
| **Delta** | B.1.617.2 | hCoV-19/Finland/THL-202117309/2021 | EPI_ISL_2557176 | MZ945494 |
| **Omicron /BA.1** | B.1.1.529 | hCoV-19/Finland/THL-202126660/2021 | EPI_ISL_8768822 | OM393712 |

**Supplementary Figure 1.** Neutralizing potency of serum, calculated as the ratio of neutralizing activity (MNT titer) to the SARS-CoV-2 anti-spike antibody concentration (binding antibody unit (BAU)/ml) specific to the receptor binding domain, RBD-IgG (A) and full-length spike protein, SFL-IgG (B)) against WT SARS-CoV-2 in uninfected subjects after two doses of COVID-19 vaccine, Comirnaty (vaccine only, n=20) and in subjects with previous SARS-CoV-2 infection followed by one dose of COVID-19 vaccine, Comirnaty (hybrid immunity, n=29). Serum samples were collected 14 to 30 days and 22 to 85 days after last vaccination for vaccine only and hybrid immunity groups, respectively. Statistical significance was determined by Mann-Whitney test, P values are indicated.

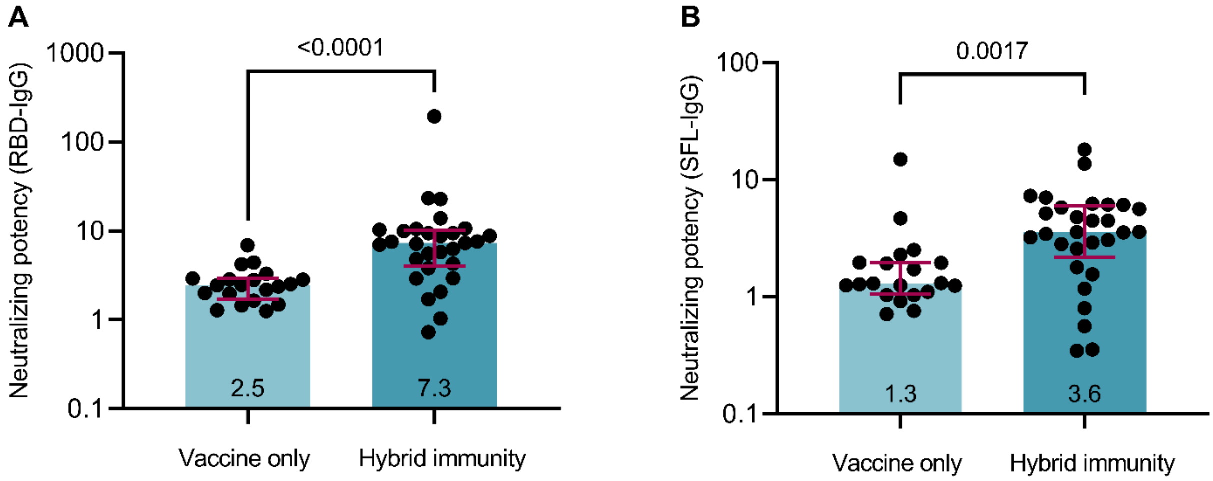
A
<0.0001
Neutralizing potency (RBD-IgG)
1000
100
10
1
0.1
2.5
7.3
Vaccine only
Hybrid immunity
B
0.0017
Neutralizing potency (SFL-IgG)
100
10
1
0.1
1.3
3.6
Vaccine only
Hybrid immunity
